# Babesia microti with multiple resistance mutations detected in an immunocompromised patient receiving atovaquone prophylaxis

**DOI:** 10.1101/2022.12.16.22283586

**Authors:** Nolan R. Holbrook, Erik H. Klontz, Gordon C. Adams, Samuel R. Schnittman, Nicolas C. Issa, Sheila A. Bond, John A. Branda, Jacob E. Lemieux

## Abstract

We report *Babesia microti* genomic sequences with multiple mutations in the atovaquone-target region of cytochrome b and the azithromycin-associated ribosomal protein L4, including newly identified mutations. The parasite was sequenced from an immunocompromised patient on prophylactic atovaquone for *Pneumocystis* pneumonia for several weeks before the diagnosis of babesiosis.

## Introduction

The intraerythrocytic parasite *Babesia microti* is the primary cause of human babesiosis in North America and is transmitted through deer tick (*Ixodes scapularis*) bites or rarely through blood transfusion^1^. While fatigue, fever, and malaise are common symptoms of a mild infection, severe babesiosis can lead to life-threatening complications primarily in older individuals and immunocompromised hosts^1,2^. The Infectious Disease Society of America recommends a seven-to-ten-day course of atovaquone and azithromycin, or alternatively, clindamycin and quinine, which is often extended for immunocompromised hosts who are at higher risk for delayed clearance and relapse^2^. Relapsing babesiosis is most frequently reported in individuals with impaired splenic function and patients with severely compromised immune systems, particularly those treated with rituximab for B-cell lymphoma^1,3–5^.

Genetic variation in *B. microti* may also be associated with clinical relapse^6^. Several previously described variants include the molecular targets of atovaquone (cytochrome b; *cytb*) and azithromycin (ribosomal protein L4; *rpl4*) that potentially confer resistance to these drugs in *B. microti*^4,6–11^. Atovaquone is known to hinder electron transport in the mitochondria of *Plasmodium* parasites at the cytochrome b site^12^, which is also the drug’s target in other apicomplexan parasites^13^. Using whole genome sequencing and target enrichment, we identified multiple mutations in the *cytb* and *rpl4* genes of *B. microti* in an immunocompromised patient who was on atovaquone prophylaxis for several weeks before babesiosis diagnosis.

### Case Report

The patient was a man in his 60s with a history of mantle cell lymphoma on rituximab (most recent dose 15 months prior to hospitalization) and acalabrutinib therapy. He was admitted to a New York hospital (2 months prior to hospitalization) for failure to thrive and possible hemophagocytic lymphohistiocytosis; he ultimately underwent splenectomy and was started on oral prednisone (40 mg daily) and oral ruxolitinib. In the month before hospitalization, the patient began taking oral atovaquone (1500 mg daily) as prophylaxis for *Pneumocystis jirovecii*. In the following month, the patient underwent an evaluation in Boston for possible myelofibrosis, where he was incidentally found to have numerous intraerythrocytic parasites on a peripheral blood smear.

On hospital day 0, a blood parasite smear (Figure 1A) identified *Babesia* infection with parasitemia of 17.6%, confirmed as *B. microti* by PCR. The patient continued taking oral atovaquone (750 mg twice daily) in addition to initiating treatment with oral azithromycin (500 mg daily) and intravenous clindamycin (600 mg every 8 hours), and oral doxycycline (100 mg every 12 hours); ruxolitinib and acalabrutinib were held on admission, and prednisone was tapered. On hospital day 1, in the setting of severe hemolysis, the patient received a red blood cell exchange transfusion with 0.25 fraction of cells remaining (FRC), with an expected reduction in parasitemia (Figure 1A). Clindamycin administration was also increased to a frequency of every 6 hours, and doxycycline was discontinued on day 3 without evidence of a concomitant tick-borne illness. There was a gradual increase in parasitemia from day 2 until day 7, until a second red blood cell exchange transfusion (FRC of 0.25) was performed on day 8. After the second exchange transfusion, the parasitemia began to trend downward. Clindamycin treatment was changed to oral administration (600 mg every 6 hours) on Day 13 and was continued with atovaquone and azithromycin following hospital discharge on Day 16. The patient continued triple-drug therapy for two months with a follow-up appointment showing a negative blood parasite smear and positive PCR. Clindamycin was subsequently discontinued in the setting of increased liver enzymes, and atovaquone and azithromycin were continued. During interval follow-up at five months, the patient was stable without clinical or blood smear evidence of recurrence.

**Figure 1.**
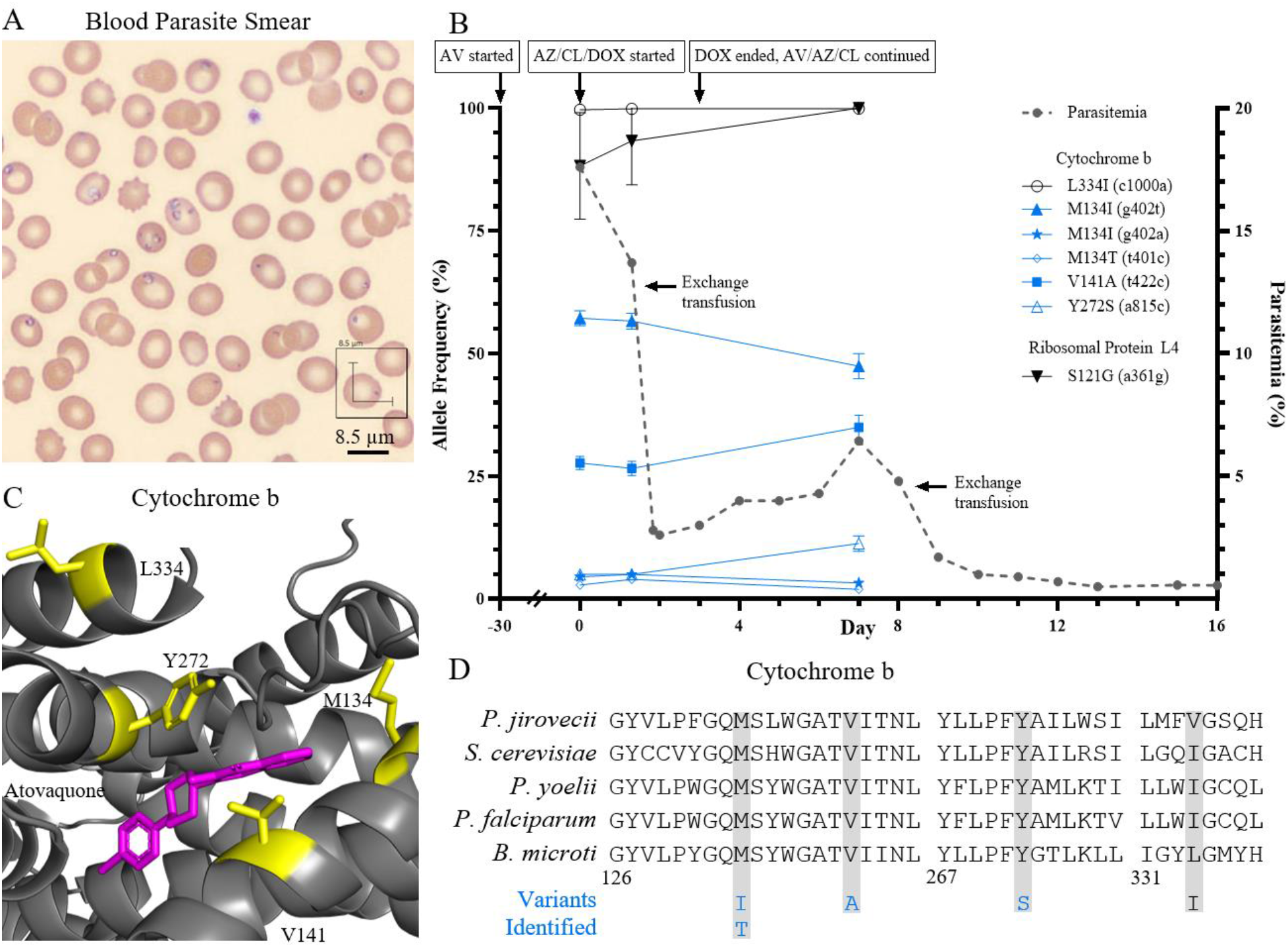
Analysis of *B. microti* clinical specimens from case report. (A) Blood parasite smear at presentation showing high-level parasitemia. (B) Clinical course of patient illness including medication dates, parasitemia percentage, and allele frequencies of variants detected, written as amino acid change with nucleotide change in parentheses. Variants predicted to cause drug resistance are colored in blue. AV, atovaquone; AZ, azithromycin; CL, clindamycin; DOX, doxycycline. The start date of atovaquone is approximate. (C) Model of *B. microti* cytochrome b with atovaquone in magenta and amino acid variants in yellow. (D) Alignment of cytochrome b from selected organisms with variants identified, numbered according to *B. microti*. Variants predicted to cause drug resistance are colored blue.

## Results and Discussion

We performed metagenomic next generation sequencing with target capture on nucleic acid derived from the patient’s blood from three time points during the first week of hospitalization, allowing us to assess changes in the allele frequency of *cytb* and *rpl4* variants (Figure 1B, Supplemental Table 1).

Within the *B. microti* mitochondrial genome, we identified six *cytb* mutations which cause amino acid substitutions in four residues of cytochrome b: Y272, V141, M134, and L334. Three of these residues (Y272, V141, and M134) are proximal to the atovaquone binding site and may decrease the parasite’s susceptibility to this drug (Figure 1C). To our knowledge, the Y272S variant has not been previously reported, but it appears similar to Y272C, which has been associated with atovaquone resistance in two cases of relapsing babesiosis^8,9^. Additionally, there is evidence that mutations in the equivalent residue (Y268) confer resistance to atovaquone in the related malarial parasite, *Plasmodium falciparum* (Figure 1D)^14–16^. We also identified a V141A substitution, which has been previously reported in a relapsing *B. microti* infection and has been suggested to confer resistance^7^. Two distinct nucleotide mutations led to the amino acid substitution M134I; at a lower allele frequency, one variant also produced M134T, which has been identified in another case^17^. These mutations underscore the importance of this position in atovaquone resistance. M134I has been previously reported in a case of relapsing babesiosis,^6^ and the equivalent mutation in *P. falciparum* results in a 25-fold increase in the IC_50_ of atovaquone,^15^ together suggesting that substitutions in M134 decrease atovaquone susceptibility in *B. microti*. Interestingly, nucleotide substitutions producing M134T, M134I, and V141A were almost exclusively found *in trans*, with allele frequencies of M134 and V141 substitutions rising and falling in opposition to each other. Under the selective pressure of atovaquone, this reciprocal pattern suggests that mutations at both sites decrease susceptibility to this drug, with the gradual rise of V141A, linked to Y272S, suggesting that this combination of mutations may confer an equivalent or even slightly greater fitness advantage in the presence of atovaquone. Finally, a novel L334I substitution was found at an allele frequency of 99.7% or higher; however, this substitution is of a very similar residue in a location outside the atovaquone binding site, making it less likely to be involved in resistance. Within the apicoplast genome, we found one variant, S121G, at high allele frequency in *rpl4*. This variant has been recently reported^7^, though its effect on azithromycin resistance is not clear, as it is not within the section of RPL4 that interacts with azithromycin in malarial models^7,18^.

In conclusion, our finding of substitutions at Y272, M134, and V141 of cytochrome b in this patient with relapsing babesiosis strengthens previous reports that mutations in these residues are associated with atovaquone resistance. We additionally report that different amino acid substitutions at these positions likely confer resistance, including the newly reported Y272S variant. The presence of these mutations is likely due to the prolonged atovaquone monotherapy used as prophylaxis for *Pneumocystis jirovecii*, which was inadvertently serving as monotherapy for *B. microti* prior to recognition and appropriate treatment of infection. The existence of multiple such mutations together in a single patient has not previously been reported and suggests the existence of more than one pathway to atovaquone resistance. Although atovaquone administration may have conferred high-level drug resistance in this variant of *B. microti*, this patient recovered after two exchange transfusions when combined with atovaquone, azithromycin, and clindamycin plus a reduction in immunosuppressive medications (prednisone, ruxolitinib, acalabruitinib). However, it is not clear why parasitemia rebounded after the first but not second exchange transfusion when no changes were made to his antimicrobial therapy; however, this might reflect the reduction in immunosuppressive medications. The use of a second exchange transfusion for the treatment of severe *B. microti* infections has been reported^19^. With a growing immunosuppressed population, antimicrobial resistance in patients with babesiosis is an increasing clinical concern. Ongoing efforts to identify mutations associated with drug resistance may inform strategies to identify drug-resistant *Babesia* more rapidly and formulate effective treatment regimens.

## Methods

### Patient Consent Statement

This work was performed under MGH/MGB-IRB protocol 2019000986, “Collection and molecular characterization of clinical excess samples tested for tick-borne illness.” Human genomic data were not analyzed in this study.

### Sequencing

Nucleotide sequences have been deposited to GenBank under accession numbers CP113809-CP113814, CP113815-CP113820, and CP113821-CP113826. Excess whole blood samples from the patient at three time points were available for analysis. DNA was extracted using the DNeasy Blood & Tissue kit (Qiagen) with 100ul of whole blood input following the manufacturer’s instructions. Sequencing libraries were prepared from the extractions using the DNA Prep kit (Illumina) following the manufacturer’s instructions. Due to areas of low coverage and read depth when sequenced directly after DNA Prep, the libraries were enriched for *B. microti* using the enrichment module from the DNA Prep with Enrichment kit (Illumina) following manufacturer recommendations for double-stranded RNA capture probes and using an updated SureSelect Custom Capture Library (Agilent) described earlier^6^, and were subsequently sequenced on a MiniSeq (Illumina). Sequencing reads were demultiplexed and aligned to the *B. microti* strain RI reference genome using viral-ngs 2.1.28 hosted on the Terra platform (app.terra.bio)^20^. Variants were identified using Geneious Prime (Dotmatics) and were included in this report if the allele fraction at any time point was >4% with at least 10 variant reads. Sections of *rpl4* (1-81 and 413-603) were not analyzed due to poor signal to noise. Graphing was performed in GraphPad Prism (Dotmatics).

### Structural modeling and alignments

The protein structure of *B. microti* cytochrome b was modeled by AlphaFold (Google) and is publicly available under UniProt entry A0A1R3Y6X4. The coordinates of atovaquone were taken from PDB accession 4PD4 after superposition of this *Saccharomyces cerevisiae* structure to *B. microti* using PyMOL (Schrӧdinger). Alignments were created in Geneious Prime using Geneious Alignment and GenBank accession numbers AFV57344.1, XP_022811206.1, UVN15258.1, CTQ41577.1, NP_009315.1.

## Data Availability

All data produced has been deposited as nucleotide sequences to GenBank under accession numbers CP113809-CP113814, CP113815-CP113820, and CP113821-CP113826.

## Acknowledgment

We thank Sanjat Kanjilal for helpful discussions and assistance with sample handling. We thank Rockib Uddin for his assistance with the hybrid capture protocol.

