## Supplemental Table 1 for "Babesia microti with multiple resistance mutations detected in an immunocompromised patient receiving atovaquone prophylaxis"

**Supplementary Data:**

|  | Day 0 | Day 1 | Day 7 |
| --- | --- | --- | --- |
| *Cytb* variants |  |  |  |
| Y272S (a815c) | 0.050 | 0.050 | 0.113 |
| V141A (t422c) | 0.277 | 0.266 | 0.350 |
| M134I (g402t) | 0.572 | 0.566 | 0.474 |
| M134I (g402a) | 0.045 | 0.050 | 0.032 |
| M134T (t401c) | 0.028 | 0.040 | 0.019 |
| L334I (c1000a) | 0.997 | 0.999 | 0.999 |
| *Rpl4* variant |  |  |  |
| S121G (a361g) | 0.882 | 0.933 | 1.000 |

Supplemental Table 1. Allele frequency of identified variants at each sample time point.
